# Analysis of the ARTIC V4 and V4.1 SARS-CoV-2 primers and their impact on the detection of Omicron BA.1 and BA.2 lineage defining mutations

**DOI:** 10.1101/2022.12.01.22282842

**Authors:** Fatima R. Ulhuq, Madhuri Barge, Kerry Falconer, Jonathan Wild, Goncalo Fernandes, Abbie Gallagher, Suzie McGinley, Ahmad Sugadol, Muhammad Tariq, Daniel Maloney, Juliet Kenicer, Rebecca Dewar, Kate Templeton, Martin McHugh

## Abstract

The ARTIC protocol uses a multiplexed PCR approach with two primer pools tiling the entire SARS-CoV-2 genome. Primer pool updates are necessary for accurate amplicon sequencing of evolving SARS-CoV-2 variants with novel mutations. The suitability of the ARTIC V4 and updated V4.1 primer scheme was assessed using whole genome sequencing of Omicron from clinical samples using Oxford Nanopore Technology. Analysis of Omicron BA.1 genomes revealed that 93.22% of clinical samples generated improved genome coverage at 50x read depth with V4.1 primers when compared to V4 primers. Additionally, the V4.1 primers improved coverage of BA.1 across amplicons 76 and 88, which resulted in the detection of the variant defining mutations G22898A, A26530G and C26577G. The Omicron BA.2 sub-variant (VUI-22JAN-01) replaced BA.1 as the dominant variant by March 2022, and analysis of 168 clinical samples showed reduced coverage across amplicons 15 and 75. Upon further interrogation of primer binding sites, a mutation at C4321T (present in 163/168, 97% of samples) was identified as a possible cause of complete dropout of amplicon 15. Furthermore, two mutations were identified within the primer binding regions for amplicon 75: A22786C (present in 90% of samples) and C22792T (present in 12.5% of samples). Together, these mutations may result in reduced coverage of amplicon 75 and further primer updates would allow the identification of the two BA.2 defining mutations present in amplicon 75; A22688G and T22679C. This work highlights the need for ongoing surveillance of primer matches as circulating variants evolve and change.

## Introduction

On the 11^th^ March 2020, the World Health Organisation (WHO) declared a global pandemic following the emergence of the severe acute respiratory syndrome coronavirus 2 (SARS-CoV-2). To date, 5^th^ September 2022, there have been over 600 million confirmed cases and over 6.5 million deaths worldwide [1].

Worldwide genomic surveillance of SARS-CoV-2 has been employed to better understand the viral mutation rate and transmission dynamics of emerging SARS-CoV-2 variants. Identifying the genetic diversity of circulating SARS-CoV-2 variants aids research into understanding the pathogenicity of variants, identifying sites on the genome that may reduce the effectiveness of vaccines and drug therapies, and ultimately inform the design of future therapeutics [2–8]. Additionally, real-time surveillance of SARS-CoV-2 outbreaks informs the implementation of control and prevention measures for SARS-CoV-2 transmission. As of 6^th^ September 2022, 12.9 million SARS-CoV-2 genomes have been submitted to the global initiative on sharing all influenza data (GISAID) database (GISAID - gisaid.org).

The most widely used protocol to sequence SARS-CoV-2 is the ARTIC amplicon-based targeted whole-genome sequencing approach. The ARTIC protocol is a freely available protocol allowing for rapid responses to changes in the SARS-CoV-2 viral genome [9–14]. This is a multiplex PCR approach, which consists of 98 pairs of primers that spans the ∼30kb genome. However, one of the major limitations of the amplicon approach is the static nature of the primer scheme as the virus evolves and develops mutations and structural variants, which can lead to primer mismatches and an inability to construct near-complete viral genomes. Since January 2020 the primer scheme has been updated four times in response to changes in the prevailing virus lineages [9–11, 13, 14]. Inability of a given primer to bind to the complementary sequence results in amplicon dropouts and mutations in the Beta, Delta and Gamma variants resulted in less efficient amplification of amplicons 72, 74 and 76 [14, 15].

The ARTIC V4.1 primers were introduced to improve primer binding and eliminate amplicon dropouts for BA.1. In this work the suitability of the ARTIC V4.1 primer scheme by comparison with the V4 primer scheme will be assessed and reported for the Omicron BA.1 lineage. To do this, Nanopore sequencing was used to sequence 70 SARS-CoV-2 samples, with both V4 and V4.1 primers, obtained from RT-PCR confirmed COVID-19 positive patients. Subsequently, BA.2 was assigned as a variant under investigation (VUI-22JAN-01) and has increased in prevalence. The ability of the V4.1 primer scheme to generate near complete genome coverage of BA.2 was also investigated in this work.

## Methods

### Patient samples and RNA extraction

344 SARS-CoV-2 positive clinical samples were collected between Dec-2021 and Mar-2022. There were 325 nose and throat swabs (94.48%), 18 throat swabs (5.23%) and 1 tracheal aspirate (0.29%). Samples were confirmed positive for at least two SARS-CoV-2 real-time RT-PCR targets using a range of diagnostic assays in NHS laboratories across the east of Scotland and referred to the Viral Sequencing Service, Royal Infirmary of Edinburgh, for sequencing. RNA extraction was performed on confirmed SARS-CoV-2 samples using two automated extraction platforms; the Biomérieux NucliSENS EMAG^®^ and Abbot *m2000sp*. First, SARS-CoV-2 positive samples were inactivated and lysed using NucliSENS^®^ easyMAG^®^ Lysis Buffer (Biomérieux, ref 280134). When using the *m2000sp*, VTM and lysis buffer were mixed at a 1:1 ratio and equal volumes of diluted lysis buffer was added to each sample for lysis (500 μl lysis buffer + 500 μl sample). For the Biomérieux NucliSens EMAG^®^, neat lysis buffer was added to each sample (200 μl sample + 2 mL lysis buffer). The procedure was followed as outlined in the manufacturer’s instructions and the RNA eluted in 110 μl. VTM only was used as an internal control for each extraction run.

### Ethical approval

This work was performed as a diagnostic lab validation and ethical approval was obtained from the NHS Lothian NRS BioResource and Tissue Governance Unit (reference 10/S1402/33).

### ARTIC LoCost protocol Nanopore library preparation

ONT libraries for SARS-CoV-2 sequencing was performed according to the ARTIC Lo-Cost protocol [11, 12], with both V4 and V4.1 primers. Slight modifications to the published method were made during the DNA barcode ligation step. For a single reaction, the ligation mastermix was prepared by adding nuclease-free water (2.1 μl), Ultra II Ligation Mastermix (5.0 μl), Ligation Enhancer (0.2μl) (NEBNext^®^ Ultra™ II Ligation Module, E7595S) and the unique barcode (1.2 μl) (ONT Native Barcoding Expansion 96, EXP-NBD196). The end prep reaction mixture (1.5 μl), from the previous step, was added to the ligation mastermix (8.50 μl) containing the unique barcodes. The protocol was then followed as outlined in the ARTIC Lo-Cost protocol [11].

### Nanopore sequence analysis

High accuracy basecalling was performed using Guppy (v4.4.0) (Oxford Nanopore Technologies) and demultiplexed using guppy_barcoder with the option ‘require_barcodes_both_ends’ and a minimum score of 60. The downstream analysis was performed using the ARTIC fieldbioinformatics pipeline (v1.2.1) (https://github.com/artic-network/fieldbioinformatics) and nanopolish variant calling (https://github.com/jts/nanopolish) to generate a consensus sequence for each sample in FASTA format. RAMPART (v1.0.6) (https://github.com/artic-network/rampart) was used to calculate genome coverage at 20x, 100x and 200x read depth. Nextclade (https://clades.nextstrain.org) was used to determine the number of SNPs between the reference genome (NC_045512.2) and sequenced clinical isolates. PANGOLIN (v3.1.20) (https://github.com/cov-lineages/pangolin) [16, 17] was used to assign a lineage to all SARS-CoV-2 samples, using the pangoLEARN algorithm. Aln2type (v0.0.3) (https://github.com/connor-lab/aln2type) (analysis performed on 8th Feb 2022) was used to identify variant defining mutations for variants of concern (VOC) and variants under investigation (VUI) as curated by Public Health England (https://github.com/phe-genomics/variant_definitions).

The metrics and results of all experiments are available in the supplementary data (Datasets 1, 2 and 3). For direct V4 to V4.1 primer comparison, two separate 48-plex libraries were loaded (Run1 and Run2, Dataset 1). For the analysis of BA.1 and BA.2 samples with V4.1 primers only, a total of 7, 48-plex libraries were loaded and sequenced.

### Sequence alignment

Sequences were aligned to the reference SARS_CoV-2 isolate NC_045512.2 using MAFFT (v7.505) (https://anaconda.org/bioconda/mafft) [18] and visualised using Jalview2 (v2.11.2.2) [19]. Depth at each nucleotide position was extracted using SAMtools (v1.15) (https://github.com/samtools/samtools).

### Quality Control

Samples were run as 48-plex libraries, each included a negative and positive control (SARS-CoV-2 B.1 lineage, collected June 2020), and two extraction negative controls (Datasets 1, 2 and 3). For all sequencing runs, the negative control and extraction negative controls generated <5 mapped reads and 0% genome coverage at 20x. The viral lysate of the positive control generated 99.90% genome coverage at 20x read depth. All seven expected mutations were detected for the positive control and assigned the PANGO lineage B.1. Clinical samples that generated <70% whole genome coverage were excluded from further analysis.

### Data availability statement

The data have been deposited with links to BioProject accession number PRJNA902683 in the NCBI BioProject database (http://www.ncbi.nlm.nih.gov/bioproject/902683).

### Conflict of interest statement

The authors have declared no conflict of interest.

## Results

### Genome coverage of Omicron BA.1 with ARTIC V4 and V4.1 primers

In total, nine (13%) clinical samples were identified as Delta (VUI-21OCT-01 or VOC-21APR-02), 59 (84%) clinical samples were identified as Omicron lineage BA.1 (VOC-21NOV-01) and two (3%) clinical samples were identified as Omicron BA.2 (VUI-22JAN-01) with ARTIC V4 and V4.1 primers (Dataset 1). For the Delta lineage samples, the median genome coverage at 20x read depth improved from 99.40% with V4 primers to 99.90% with V4.1 primers, respectively. All nine samples were confirmed as either VUI-21OCT-01 or VOC-21APR-02 with both V4 and V4.1 primers. The two Omicron BA.2 samples increased from 95.20% to 99.30% and 97.20% to 99.90% genome coverage, with V4 and V4.1 primers, respectively. Both samples had probable aln2type VUI-22JAN-01 calls with V4 primers, however one sample improved from probable to confirmed with V4.1 primers.

Initial in-depth analysis focussed on Omicron BA.1, as the increasingly dominant variant and the main component of the runs during sequencing. The genome coverage at >50x read depth was investigated to identify regions of the genome with reduced amplification. Nucleotide positions that generated <50x read depth in 15% or more of the samples were considered as low coverage regions. Coverage was compared between the V4 and V4.1 primer scheme. The V4.1 primer scheme included a new batch of primers added to the V4 primers to improve the coverage across amplicons 10, 23, 27, 76, 79, 88, 89 and 90 [13].

Use of the V4 primers generated reduced amplification of nucleotides across 18 different amplicons, with less than 50x read depth in more than 15% of sequenced samples (Figure 1A, Table S1). The use of the V4 primers resulted in dropout of amplicons 76 and 90 (Figures 1A and 2A, Table S1) and poor coverage across amplicons 10, 23, 79, 88 and 89 (Figures 1A and 2A, Table S1). Reduced coverage of these amplicons was due to primer mismatches between the V4 primer scheme and the SARS-CoV-2 BA.1 sequence. Reduced coverage of nucleotide positions across amplicons 22, 37, 72, 73, 74 and 95 were due to small deletions within the genome (3-9bp) (Table S1). Reduced amplification of amplicons 21, 29, 31, 51 and 60 (Figure 1A, Table S1) were detected with the V4 primers in samples that generated <60,000 mapped reads. Due to the low number of mapped reads for these samples, the reduced coverage within these amplicons is likely to be due to the quality and quantity of the extracted RNA.

**Figure 1.**
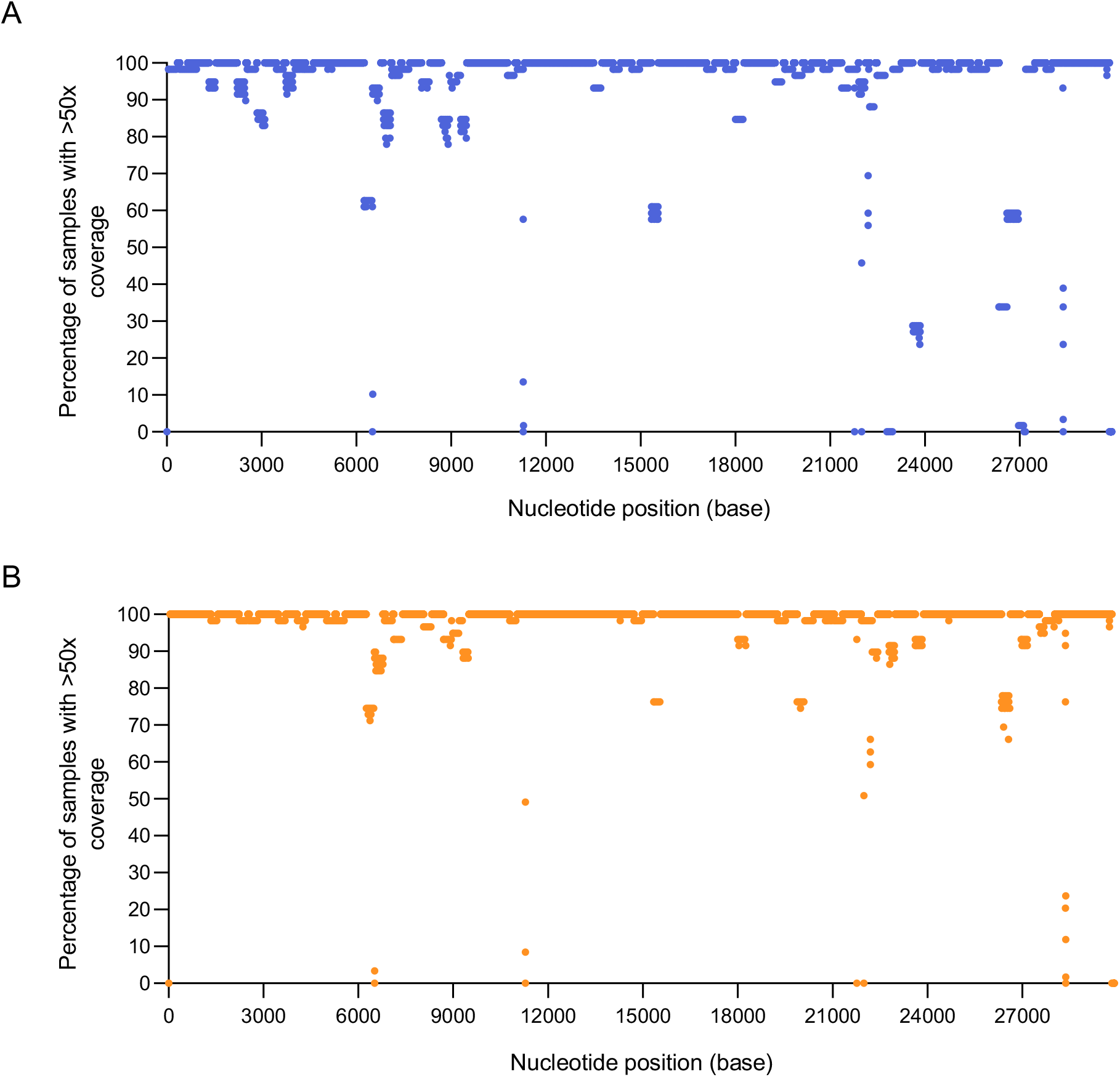
Coverage profile of SARS-CoV-2 BA.1 using V4 and V4.1 primers. Percentage of samples with >50x coverage at each nucleotide position for the assembly of 59 SARS-CoV-2 BA.1 samples with **A**. V4 primers (in blue) or **B**. V4.1 primers (in orange). The depth of each nucleotide position was generated using SAMtools on the primer trimmed alignment files.

Utilisation of the V4.1 primer scheme [13] resulted in an overall improvement in genome coverage. The addition of the V4.1 alt primers increased the median sequencing depth across all eight amplicons with the modified primers in the updated V4.1 primer scheme (Figure 2A). Strikingly, the additional primers rescued the dropout of amplicons 76 and 90 and increased sequencing coverage across amplicons 10, 23, 78, 88 and 89 (Figure 2A). The number of amplicons with nucleotide positions that generated <50x coverage in 15% or more samples decreased from 18 amplicons with V4, to 10 amplicons with V4.1 primers (Figure 1B, Table S2). Despite the addition of modified primers to better amplify amplicon 88, the coverage defect was not completely restored. However, the percentage of sequenced samples <50x coverage within this region decreased, from 40.68 - 66.1% to 22.03 - 33.9%, with V4 and V4.1 primers, respectively (Tables S1 and S2). Regions with lower coverage with the V4.1 primers were investigated to identify the root cause of lower read depth at certain nucleotide positions. The reduced coverage across amplicons 22, 37, 72, 73, 74 and 95 were due to small deletions within the genome (3-9 bp). Reduced amplification of amplicons 21, 51 and 66 was detected in samples that generated <60,000 mapped reads, suggesting overall lower genome quality.

**Figure 2.**
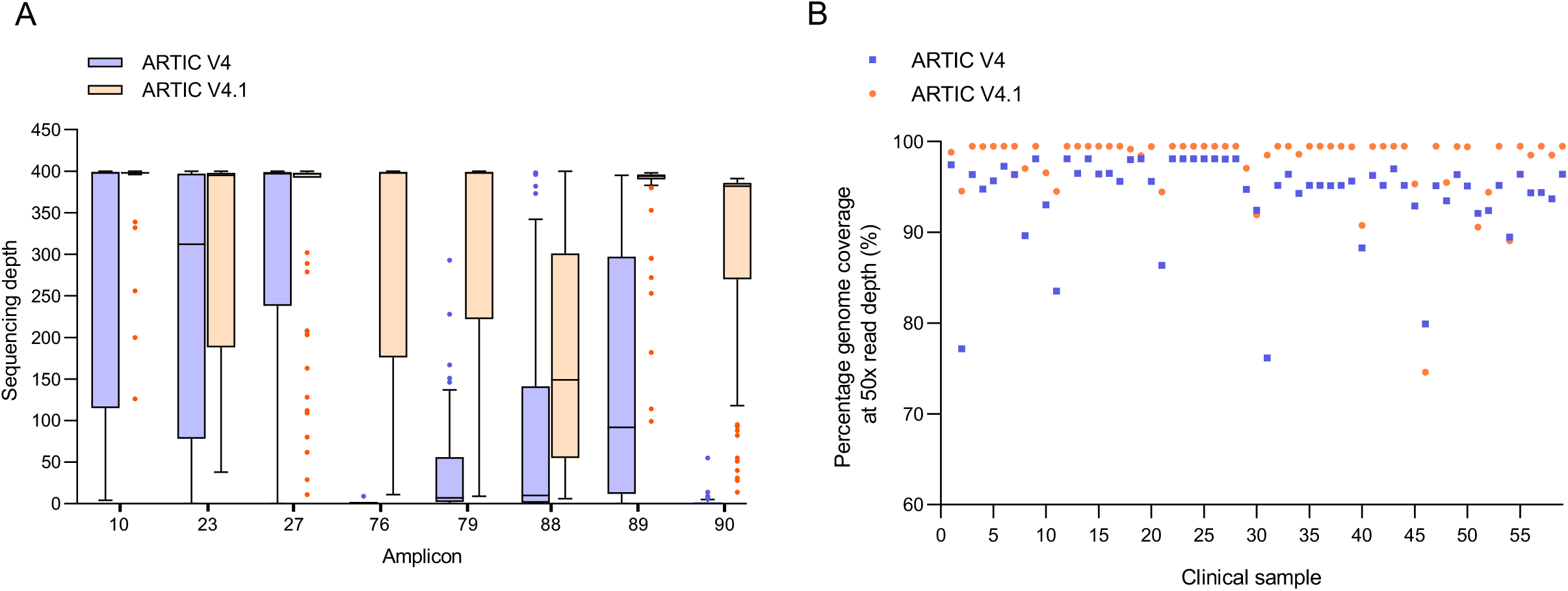
Increased sequencing depth across amplicons and increased genome coverage in SARS-CoV-2 BA.1 samples amplified with V4.1 primers. **A**. Median coverage across amplicons 10, 23, 27, 76, 79, 88, 89 and 90 with V4 (blue) and V4.1 (orange). **B**. Percentage genome coverage at >50x coverage of all 59 SARS-CoV-2 BA.1 samples. Assemblies with V4 primers in blue, and V4.1 primers in orange.

Analysis of all BA.1 sequences highlighted that in total, 2668 nucleotides generated reduced coverage (<50x read depth in 15% or more samples) with the V4 primers compared to 1085 nucleotides with the V4.1 primers (Tables S1 and S2). The median number of reads mapped per sample was 98,609 and 114,476 with V4 and V4.1 primers, respectively (Dataset 1). Additionally, the median number of masked bases (N’s) decreased from 1050 N’s to 126 N’s with the V4 and V4.1 primers, respectively (Dataset 1). In addition, the median number of SNPs increased from 52 to 58 with the V4 and V4.1 primers, respectively. This highlights the overall improvement detected with the V4.1 primers when compared to V4. The percentage of samples that generated improved genome coverage with the V4.1 primers when compared to V4 was 93.22% (55/59) at 50x read depth (Figure 2B). Additionally, 37 out of 59 (62.71%) samples generated increased number of mapped reads with the V4.1 primers compared to V4 (Dataset 1). The median genome coverage per sample for V4 primers at 200x, 100x and 20x was 95.59%, 96.30% and 98.00%, respectively (Dataset 1). The median coverage per genome with V4.1 primers at 200x, 100x and 20x was 98.80%, 99.60%, and 99.90%, respectively.

### PANGO Lineage and VOC/VUI calls of Omicron BA.1 with ARTIC V4 and V4.1 primers

PANGOLIN was used to assign a lineage to all SARS-CoV-2 consensus sequences and 58/59 samples received identical PANGO lineages, either BA.1 (37/59) or BA.1.1 (21/59) with V4 and V4.1 primers (Dataset 1). The specificity of the lineage call for vssfru_032 altered from BA.1 to BA.1.1 with the V4.1 primers (vssfru_032b) when compared to V4 (vssfru_032b) (Dataset 1). Based on PHE variant definitions, aln2type requires the detection of 17 variant defining mutations to be assigned as SARS-CoV-2 BA.1 (VOC-21NOV-01). All 17 variant defining mutations must be detected for a ‘confirmed’ VOC/VUI status call. Interestingly, more accurate VOC-21NOV-01 status calls were obtained in 55/59 samples (93.22%), improving from probable to confirmed VOC-21NOV-01 when using V4.1 compared to V4 primers (Dataset 1). The remaining 4/59 samples (6.78%) had probable VOC-21NOC-01 calls with both V4 and V4.1 primers (Dataset 1).

Further analysis highlighted that the median number of variant defining mutations detected for VOC-21NOV-01, increased from 14 to 17 with the V4 and V4.1 primers, respectively. The increased coverage across amplicons 76 and 88 with the V4.1 primers (Figure 2A) resulted in increased coverage at position 22898, 26530 and 26577 (Figure 3A) and the detection of the three additional variant defining SNPs, G22898A, A26530G and C26577G, present in amplicons 76 and 88 (Figure 3B). In addition, increased coverage of amplicon 60 allowed the identification of the variant defining mutation A18163G (Figure 3A and 3B). In total, 138 variant defining bases were masked (N’s) with the V4 primers and this was reduced to 12 bases with V4.1 primers (Figure 3B). The 12 masked variant defining bases at position 18163 (n=2), 22898 (n=2), 26530 (n=4) and 26577 (n=4) were only detected in 4 BA.1 samples (Figure 3B) that generated <30,000 mapped reads, therefore overall reduced genome quality.

**Figure 3.**
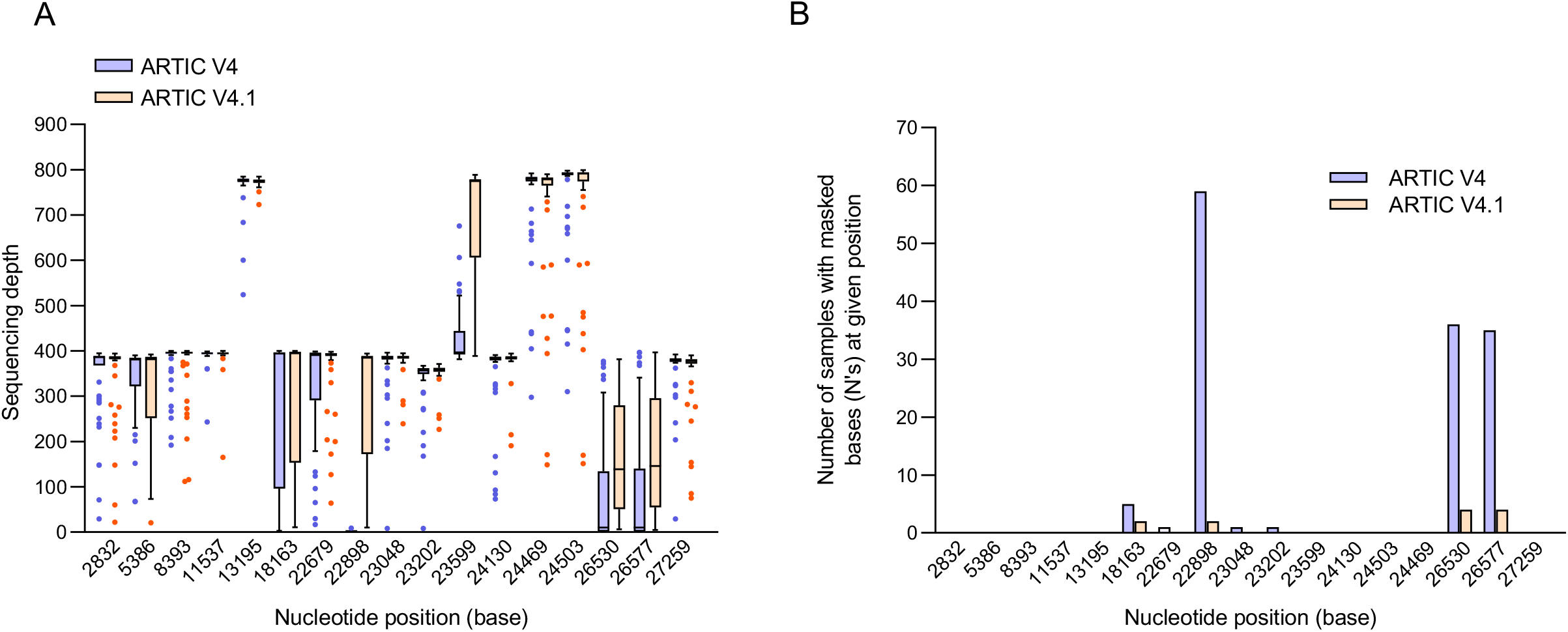
Increased sequencing depth at BA.1 variant defining positions 22898, 26530 and 26577. Analysis of 59 BA.1 samples with V4 and V4.1 primers. **A**. Sequencing depth of variant defining mutations at all 17 nucleotide positions as defined by aln2type for BA.1 with V4 (blue) and V4.1 (orange) primers. **B**. Number of samples with masked bases (N’s) at variant defining nucleotide positions in BA.1 samples with V4 (blue) and V4.1 (orange) primers.

A larger dataset of 106 positive SAR-CoV-2 BA.1 samples collected between 21-02-2022 and 07-03-2022 were investigated to compare the aln2type results obtained from the side-by-side V4 and V4.1 comparison above. Comparable results were obtained with reduced coverage at positions 18163, 22898, 26530 and 26577 (present in amplicons 60, 76 and 88) with the V4.1 primers (Figure S1A and S1B). Reduced amplification of these regions contributed to only 16/106 (15.1%) of samples with probable VOC-21NOV-01 calls (Dataset 2) and a total of 48 masked variant defining mutations (Figure S1B, Dataset 2). Altogether, these results highlight the overall improvement in the amplification of Omicron BA.1 with the V4.1 primers when compared to V4.

### Genome coverage of Omicron BA.2 with ARTIC V4.1 primers

Genome coverage of Omicron BA.2 (VUI-22JAN-01) was further assessed with the ARTIC V4.1 primers by analysing 168 SARS-CoV-2 positive BA.2 samples collected from 2022-02-21 – 2022-03-06. Overall, the median genome coverage of 168 BA.2 samples with V4.1 primers at 200x, 100x and 20x were 96.15%, 98.60% and 99.30%, respectively (Dataset 3). The median number of mapped reads was 82,598 (Dataset 3). The median number of masked bases (N’s) was 387 with a median number of 68 SNPs across the BA.2 genomes. In total, 16 amplicons were identified with <50x read depth in more than 15% of the samples sequenced (Figure 4, Table S3). Reduced coverage of nucleotides across amplicons 37, 72, 95 and 99 were due to deletions within BA.2 (8-26 bp deletions). It was not possible to identify a cause for the reduced read depth of nucleotides present in amplicons 1, 21, 22, 51, 60, 66, 74, 76, 88 or 90, however, BA.2 samples with reduced nucleotide coverage in these regions generated a total of <60,000 mapped reads per genome. This suggests the low coverage may be due to pre-analytical issues with the amount and quality of RNA. Dropout of amplicon 15 and poor coverage across amplicon 75 was detected (Figure 4, Table 3). BA.2 samples with reduced nucleotide coverage within these regions generated >60,000 mapped reads. These samples were further investigated to identify the root cause of reduced coverage.

**Figure 4.**
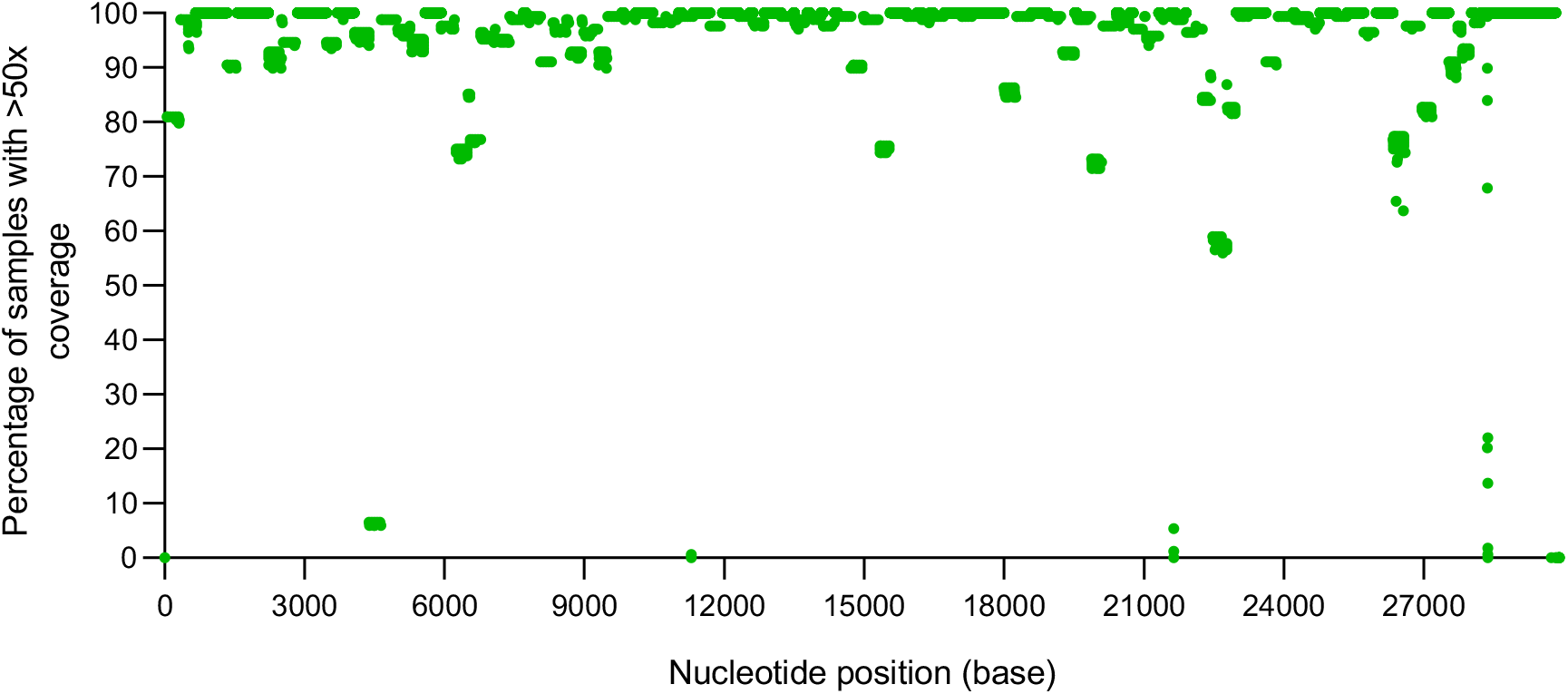
Coverage profile of SARS-CoV-2 BA.2 using V4.1 primers. Percentage of samples with >50x coverage at each nucleotide position for the assembly of 168 SARS-CoV-2 BA.2 samples with V4.1 primers (in green). The depth of each nucleotide position was generated using SAMtools on the primer trimmed alignment files.

With regards to amplicon 15, a SNP (C4321T) was identified in 163/168 samples (97%) (Figure 5A). The primer ‘SARS-CoV-2_15_LEFT’ binds within this region of the genome and the C4321T SNP sits 9 bp into the forward primer (Figure 5A). In amplicon 75 two SNPs were identified within the primer binding region of ‘SARS-CoV-2_75_RIGHT’, A22786C and C22792T (Figure 5B). The variant defining mutation, A22786C is present in 90% of samples (151/168) while C22792T is present in 12.5% of samples (21/168). The variant defining mutation, A22786C sits at the end of the ‘SARS-CoV-2_75_RIGHT’ primer (3’ region), therefore does not have a significant impact on primer binding. However, samples with the C22792T SNP exhibit dropouts in amplicon 75, highlighting the importance of this site for primer binding (Figure 5B). Updates to these primers may provide improved coverage of amplicons 15 and 75.

**Figure 5.**
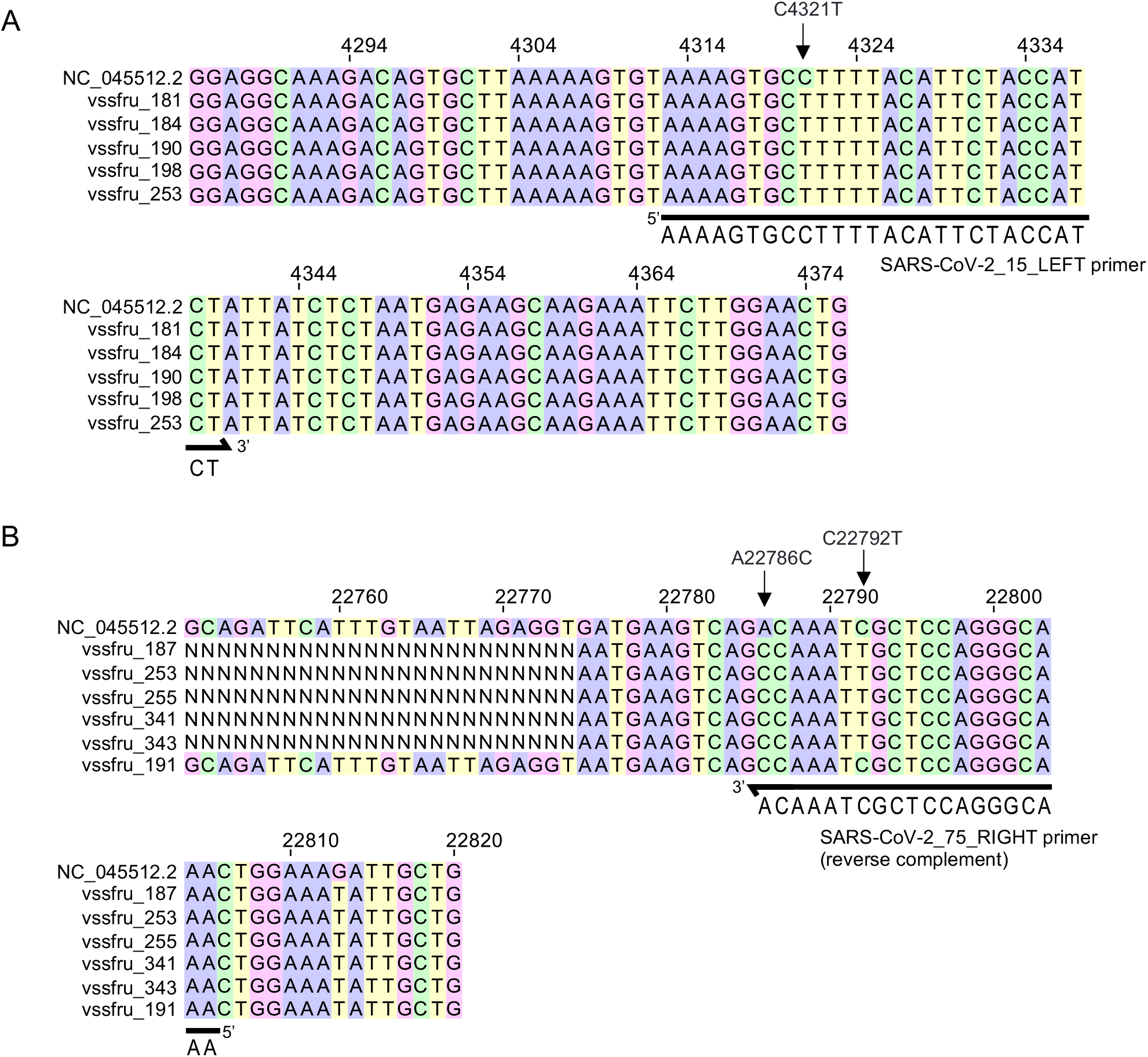
Identification of SNPs responsible for low coverage of amplicons 15 and 75 in BA.2 samples. Multiple sequence alignment using MAFFT (v7.505) of NC_045512.2 reference with selected BA.2 samples with **A**. reduced coverage of amplicon 15 or **B**. reduced coverage of amplicon 75. Alignments viewed using Jalview (v2.11.2.2).

Reduced amplification of amplicons 75, 76 and 88 resulted in reduced sequencing coverage of four variant defining SNPs (T22679C, A22688G, A22786C and C26577G) (Figure S2, Table S3) required for aln2type Omicron BA.2 variant under investigation (VUI-22JAN-01) call criteria (Table S3). BA.2 (VUI-22JAN-01) required detection of 20 variant defining mutations for a confirmed status call and 80 probable VUI-22JAN-01 calls were obtained with a total of 197 masked bases (N’s) at variant defining positions across all BA.2 samples sequenced (Figure S2B, Dataset 3). Of these, 71 BA.2 samples generated over 90% genome coverage at 20x read depth and were then further investigated. The 71 probable VUI-22JAN-01 samples accumulated a total of 131 masked bases (N’s) at the following variant defining positions: 2790 (n=3), 9424 (n=29), 18163 (n=7), 22679 (n=34), 22688 (n=34), 22775 (n=5), 22786 (n=9), 26577 (n=10) (Dataset 3). As expected, masked bases (N’s) were detected at position 22679, 22688, 22786 and 26577 due to lower coverage across amplicons 75, 76 and 88. However, the number of samples with masked bases at position 9424 (n=29) was unexpected, as 27 out of the 29 samples generated more than 20x coverage at position 9424 (Table S4). Further analysis revealed that the discrepancy in the detection of A9424G was due to the presence of mixed bases, with variant base calls often having low quality. This resulted in masking of the variant defining SNP A9424G in our consensus calling pipeline due to low support for the variant call.

In addition, reference base calls at variant defining sites, at either position 670 (6/168, 4%) or 9866 (1/168, 0.6%) (Table S5) contributed to probable VUI-22JAN-01 calls in seven BA.2 samples. This might be due to diversity within the BA.2 population, where reversion mutations may have occurred. Furthermore, a small cluster of 10 BA.2 samples were defined as having mixed calls for the variant defining mutation at position 29510 (Table S5). An alignment comparing vssfru_253 (confirmed A29510C SNP), the SARS-CoV-2 reference genome (NC_045512.2) and the 10 BA.2 samples with aln2type mixed call status (Figure S3) was generated (Figure S3). The alignment highlights the presence of A29510C within all 10 BA.2 samples, however an additional SNP, G29511T was present downstream (Figure S3). The expected codon is CGT, however, a subset of BA.2 samples have CTT within their consensus sequence, this is then defined as a mixed call by aln2type. Strikingly, the presence of this unique mutation, not identified within any other sequences analysed within this work, may indicate transmission during a small outbreak.

## Conclusion

In summary, use of the ARTIC V4.1 primers, when compared to V4 primers, improved overall genome coverage of Omicron BA.1. The addition of the alt primers improved the sequencing depth across all eight amplicons which resulted in more accurate PANGO lineage and VOC/VUI calls. Further analysis of the BA.2 sub-variant revealed complete drop-out of amplicon 15 and reduced coverage of amplicon 75. Additionally, lower read depth of amplicon 75 resulted in the masking of two variant defining mutations. This work highlights the need for ongoing surveillance of primer matches as circulating variants evolve and change. The use of different primer schemes should be an important consideration when identifying and assigning variants of concern and the genome completeness of SARS-CoV-2 genomes should be routinely monitored with updates to primer schemes when required.

## Acknowledgements

We would like to thank diagnostic laboratory teams at NHS Borders, NHS Fife, NHS Grampian, NHS Highland, NHS Lothian, NHS Orkney, NHS Shetland, and NHS Tayside for providing samples used in this work. We are also grateful to colleagues in Public Health Microbiology at Public Health Scotland, NHS Greater Glasgow and Clyde, National Services Division of NHS National Services Scotland, and Scottish Government with whom the sequencing service was established.

## Figure legends

**Figure S1.**
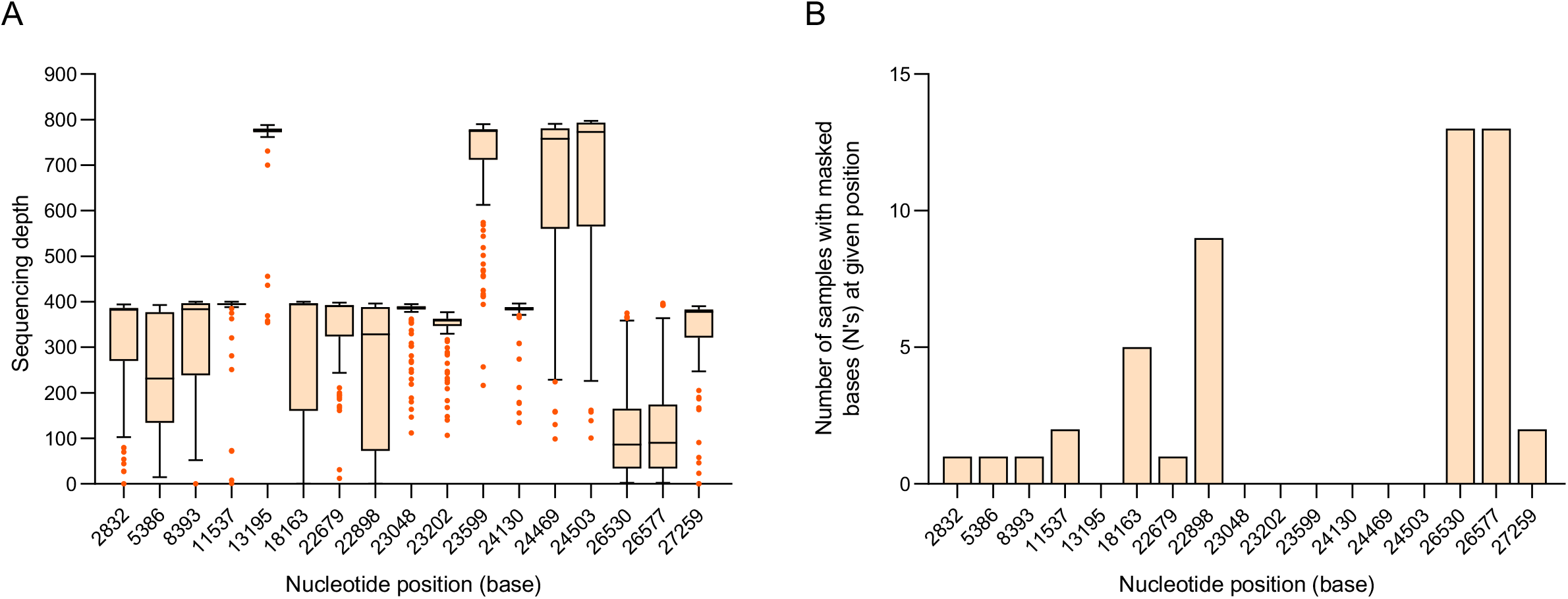
Sequencing coverage and aln2type VOC-21NOV-01 defining mutations. Analysis of 106 BA.1 samples with V4.1 primers. **A**. Sequencing depth of variant defining mutations at all 17 nucleotide positions as defined by aln2type for BA.1. **B**. Number of samples with masked bases (N’s) at variant defining nucleotide positions in BA.1 samples.

**Figure S2.**
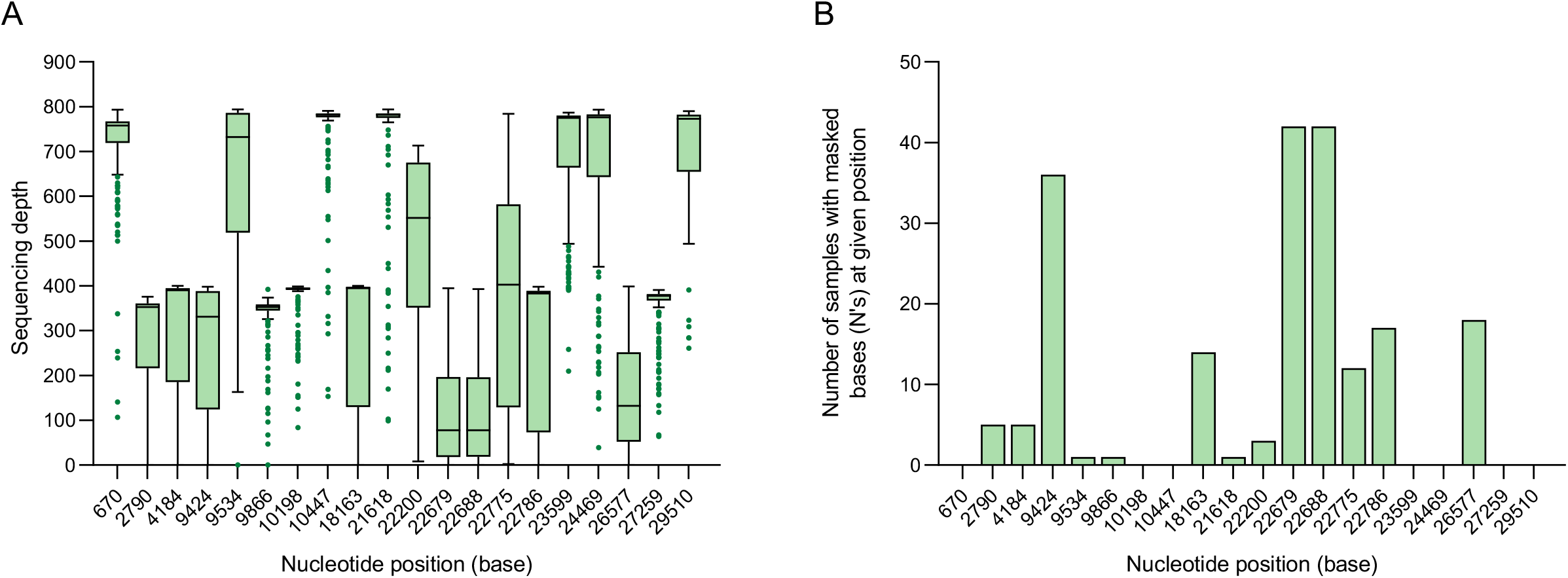
Sequencing coverage and aln2type VUI-22JAN-01 defining mutations. Analysis of 168 BA.2 samples. **A**. Sequencing depth of variant defining mutations at all 20 nucleotide positions as defined by aln2type for BA.2. **B**. Number of samples with masked bases (N’s) at variant defining nucleotide positions in BA.2 samples.

**Figure S3.**
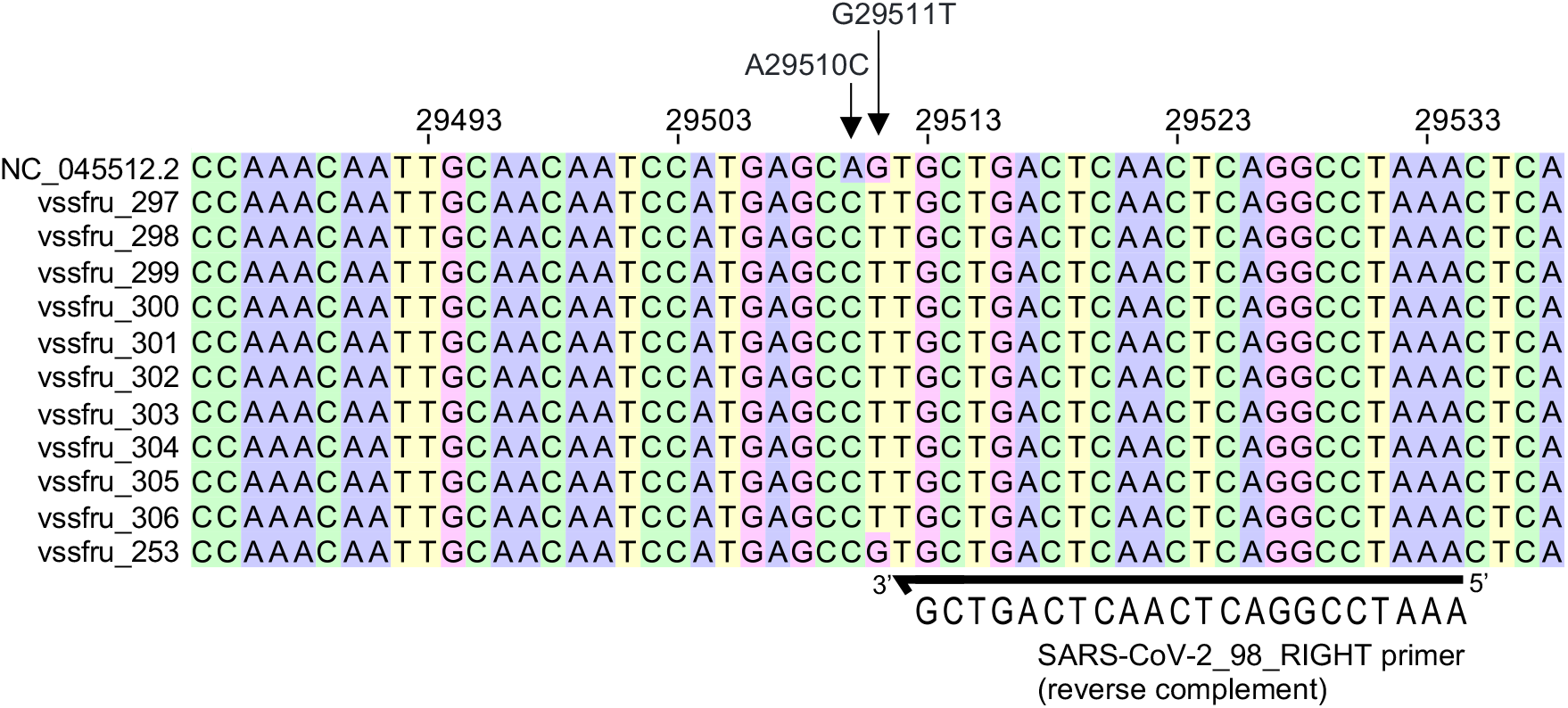
A29510C aln2type mixed-call status due to downstream mutation present in subset of samples. Multiple sequence alignment using MAFFT (v7.505) of NC_045512.2 reference with 10 BA.2 samples with mixed-call status for A29510C variant defining mutation and vssfru_253 (A29510C detected). Alignment viewed using Jalview (v2.11.2.2).

## Supplementary Datasets

**Dataset 1. Overview of SARS-COV-2 genome coverage with V4 and V4.1 primers**. Total number of SNPs and masked bases (N’s) generated using NextClade. Mapped reads, 20x, 100x and 200x coverage data generated using RAMPART (v1.0.6). Lineage and conflict generated using PANGOLIN (v3.1.20). PHE-label, status and mutation calls generated using aln2type (v0.0.3). Data from two 48-plex libraries (Run1 and 2).

**Dataset 2. Overview of SARS-COV-2 BA.1 genome coverage with V4.1 primers**. Total number of SNPs and masked bases (N’s) generated using NextClade. Mapped reads, 20x, 100x and 200x coverage data generated using RAMPART (v1.0.6). Lineage and conflict generated using PANGOLIN (v3.1.20). PHE-label, status and mutation calls generated using aln2type (v0.0.3). Data from seven 48-plex libraries (Run3 – Run9) displaying only the BA.1 samples.

**Dataset 3. Overview of SARS-COV-2 BA.2 genome coverage with V4.1 primers**. Total number of SNPs and masked bases (N’s) generated using NextClade. Mapped reads, 20x, 100x and 200x coverage data generated using RAMPART (v1.0.6). Lineage and conflict generated using PANGOLIN (v3.1.20). PHE-label, status and mutation calls generated using aln2type (v0.0.3). Data from seven 48-plex libraries (Run3 – Run9) displaying only the BA.2 samples.

## Notes

### Competing Interest Statement

The authors have declared no competing interest.

### Funding Statement

This work was performed as a diagnostic lab validation as part of the sequencing service established by the Scottish Government.

## References

1. WHO Coronavirus (COVID-19) Dashboard. https://covid19.who.int.

2. Nadeau S, Beckmann C, Topolsky I, Vaughan T, Hodcroft E, et al. Quantifying SARS-CoV-2 spread in Switzerland based on genomic sequencing data. medRxiv 2020.

3. Martin J, Klapsa D, Wilton T, Zambon M, Bentley E, et al. Tracking SARS-CoV-2 in Sewage: Evidence of Changes in Virus Variant Predominance during COVID-19 Pandemic. Viruses;12.

4. Volz E, Mishra S, Chand M, Barrett JC, Johnson R, et al. Transmission of SARS-CoV-2 Lineage B.1.1.7 in England: Insights from linking epidemiological and genetic data. medRxiv 2021.

5. Chen C, Nadeau S, Yared M, Voinov P, Xie N, et al. CoV-Spectrum: Analysis of Globally Shared SARS-CoV-2 Data to Identify and Characterize New Variants. Bioinformatics 2021;38:1735–1737.

6. Gram MA, Emborg H-D, Schelde AB, Friis NU, Nielsen KF, et al. Vaccine effectiveness against SARS-CoV-2 infection or COVID-19 hospitalization with the Alpha, Delta, or Omicron SARS-CoV-2 variant: A nationwide Danish cohort study. PLOS Med;19.

7. Andrews N, Stowe J, Kirsebom F, Toffa S, Rickeard T, et al. Covid-19 Vaccine Effectiveness against the Omicron (B.1.1.529) Variant. N Engl J Med 2022;386:1532– 1546.

8. Kirsebom FCM, Andrews N, Stowe J, Toffa S, Sachdeva R, et al. COVID-19 vaccine effectiveness against the omicron (BA.2) variant in England. Lancet Infect Dis 2022;22:931–933.

9. Quick J. nCoV-2019 sequencing protocol V.1. protocols.io.

10. Quick J. nCoV-2019 sequencing protocol v2 (GunIt) V.2. protocols.io.

11. Quick J. nCoV-2019 sequencing protocol v3 (LoCost). protocols.io.

12. Tyson JR, James P, Stoddart D, Sparks N, Wickenhagen A, et al. Improvements to the ARTIC multiplex PCR method for SARS-CoV-2 genome sequencing using nanopore. bioRxiv 2020.

13. ARTIC Network. SARS-CoV-2 V4.1 update for Omicron variant. https://community.artic.network/t/sars-cov-2-v4-1-update-for-omicron-variant/342 (2021).

14. ARTIC Network. SARS-CoV-2 version 4 scheme release. https://community.artic.network/t/sars-cov-2-version-4-scheme-release/312.

15. Davis JJ, Long SW, Christensen PA, Olsen RJ, Olson R, et al. Analysis of the ARTIC Version 3 and Version 4 SARS-CoV-2 Primers and Their Impact on the Detection of the G142D Amino Acid Substitution in the Spike Protein. Microbiol Spectr;9.

16. O’Toole Á, Scher E, Underwood A, Jackson B, Hill V, et al. Assignment of epidemiological lineages in an emerging pandemic using the pangolin tool. Virus Evol;7.

17. Rambaut A, Holmes EC, O’Toole Á, Hill V, McCrone JT, et al. A dynamic nomenclature proposal for SARS-CoV-2 lineages to assist genomic epidemiology. Nat Microbiol 2020;5:1403–1407.

18. Katoh K, Misawa K, Kuma K, Miyata T. MAFFT: a novel method for rapid multiple sequence alignment based on fast Fourier transform. Nucleic Acids Res 2002;30:3059–3066.

19. Waterhouse AM, Procter JB, Martin DMA, Clamp M, Barton GJ. Jalview Version 2--a multiple sequence alignment editor and analysis workbench. Bioinformatics 2009;25:1189–1191.

