## supplementary tables for "Analysis of the ARTIC V4 and V4.1 SARS-CoV-2 primers and their impact on the detection of Omicron BA.1 and BA.2 lineage defining mutations"

**Table S1.** Regions of the genome with <50x coverage in 15% or more BA.1 samples sequenced using V4 primers.

| Amplicon | Nucleotides with <50x read depth (n) | Range of genomes with <50x read depth across nucleotide positions (%) | VOC-21NOV-01 defining mutations affected |
|---|---|---|---|
| 10 | 68 | 15.25 – 16.95% | NA |
| 21 | 260 | 37.29 – 38.98% | NA |
| 22 | 3 | 89.83 – 100% | NA |
| 23 | 77 | 15.25 – 22.03% | NA |
| 29 | 253 | 15.25 – 22.03% | NA |
| 31 | 191 | 15.25 – 20.34% | NA |
| 37 | 9 | 42.37 – 100% | NA |
| 51 | 221 | 38.98 – 42.37% | NA |
| 60 | 270 | 15.25% | A18163G |
| 72 | 5 | 100% | NA |
| 73 | 9 | 54.24 – 100% | NA |
| 74 | 3 | 30.51 – 44.07% | NA |
| 76 | 189 | 100% | G22898A |
| 79 | 265 | 71.19 – 76.27% | NA |
| 88 | 283 | 40.68 – 66.1% | A26530G<br>C26577G |
| 89 | 274 | 40.68 – 42.37% | NA |
| 90 | 282 | 40.68 – 100% | NA |
| 95 | 6 | 61.02 – 100% | NA |

**Table S2.** Regions of the genome with <50x coverage in 15% or more BA.1 samples sequenced using V4.1 primers.

| Amplicon | Nucleotides with <50x read depth (n) | Range of genomes with <50x read depth across nucleotide positions (%) | VOC-21NOV-01 defining mutations affected |
|---|---|---|---|
| 21 | 260 | 25.42% – 28.8% | NA |
| 22 | 48 | 15.25 – 100 % | NA |
| 37 | 9 | 50.85 – 100% | NA |
| 51 | 221 | 23.73% | NA |
| 66 | 240 | 23.73 – 25.42% | NA |
| 72 | 5 | 100% | NA |
| 73 | 9 | 49.15 – 100% | NA |
| 74 | 3 | 33.9 – 40.68% | NA |
| 88 | 283 | 22.03 – 33.9% | A26530G<br>C26577G |
| 95 | 7 | 23.73 – 100% | NA |

**Table S3.** Regions of the genome with <50x coverage in 15% or more BA.2 samples sequenced using V4.1 primers.

| Amplicon | Nucleotides with <50x read depth (n) | Range of genomes with <50x read depth across nucleotide positions (%) | VUI-22JAN-01 defining mutations masked |
|---|---|---|---|
| 1 | 294 | 19.05 – 20.24% | NA |
| 15 | 261 | 93.45 – 94.05% | NA |
| 21 | 260 | 25 – 27.79% | NA |
| 22 | 229 | 15.48 – 23.81% | NA |
| 37 | 9 | 99.40 – 100% | NA |
| 51 | 221 | 24.40 – 25.60% | NA |
| 60 | 6 | 15.48% | NA |
| 66 | 240 | 26.79 – 28.57% | NA |
| 72 | 9 | 94.64 – 100% | NA |
| 74 | 181 | 15.48 – 16.07% | NA |
| 75 | 300 | 41.07 – 44.05% | T22679C<br>A22688G |
| 76 | 189 | 17.26 – 18.45% | A22786C |
| 88 | 283 | 22.62 – 36.31% | C26577G |
| 90 | 221 | 17.26 – 19.05% | NA |
| 95 | 8 | 16.07 – 100% | NA |
| 99 | 26 | 100% | NA |

**Table S4.** Sequencing coverage of VUI-22JAN-01 samples with a masked base (no-call) at position A9424G (Dataset 3).

| Sample ID | Run no. | Barcode | Position | Reference base reads | Variant base reads | Status |
|---|---|---|---|---|---|---|
| vssfru_177 | Run3 | barcode01 | A9424G | 5 | 43 | no-call |
| vssfru_179 | Run3 | barcode14 | A9424G | 3 | 15 | no-call |
| vssfru_182 | Run3 | barcode20 | A9424G | 36 | 346 | no-call |
| vssfru_184 | Run3 | barcode30 | A9424G | 31 | 255 | no-call |
| vssfru_186 | Run3 | barcode33 | A9424G | 11 | 92 | no-call |
| vssfru_189 | Run3 | barcode38 | A9424G | 28 | 211 | no-call |
| vssfru_190 | Run3 | barcode39 | A9424G | 15 | 199 | no-call |
| vssfru_192 | Run3 | barcode45 | A9424G | 40 | 350 | no-call |
| vssfru_193 | Run4 | barcode49 | A9424G | 2 | 45 | no-call |
| vssfru_194 | Run4 | barcode51 | A9424G | 4 | 40 | no-call |
| vssfru_221 | Run5 | barcode23 | A9424G | 2 | 64 | no-call |
| vssfru_233 | Run6 | barcode53 | A9424G | 1 | 9 | no-call |
| vssfru_236 | Run6 | barcode59 | A9424G | 4 | 75 | no-call |
| vssfru_239 | Run6 | barcode75 | A9424G | 7 | 47 | no-call |
| vssfru_241 | Run6 | barcode79 | A9424G | 23 | 155 | no-call |
| vssfru_247 | Run6 | barcode92 | A9424G | 43 | 334 | no-call |
| vssfru_249 | Run7 | barcode01 | A9424G | 0 | 50 | no-call |
| vssfru_257 | Run7 | barcode11 | A9424G | 45 | 344 | no-call |
| vssfru_258 | Run7 | barcode12 | A9424G | 4 | 75 | no-call |
| vssfru_259 | Run7 | barcode13 | A9424G | 23 | 183 | no-call |
| vssfru_260 | Run7 | barcode14 | A9424G | 23 | 364 | no-call |
| vssfru_266 | Run7 | barcode25 | A9424G | 19 | 364 | no-call |
| vssfru_270 | Run7 | barcode31 | A9424G | 22 | 240 | no-call |
| vssfru_272 | Run7 | barcode35 | A9424G | 29 | 360 | no-call |
| vssfru_274 | Run7 | barcode37 | A9424G | 49 | 332 | no-call |
| vssfru_280 | Run7 | barcode48 | A9424G | 18 | 188 | no-call |
| vssfru_299 | Run8 | barcode72 | A9424G | 11 | 55 | no-call |
| vssfru_308 | Run8 | barcode84 | A9424G | 5 | 57 | no-call |
| vssfru_337 | Run9 | barcode37 | A9424G | 29 | 267 | no-call |

**Table S5.** Sequencing coverage of VUI-22JAN-01 samples with 1 reference call at a variant defining position as defined by aln2type (no-detect) (Dataset 3).

| Sample ID | Run no. | Barcode | Position | Reference base reads | Variant base reads | Status |
|---|---|---|---|---|---|---|
| vssfru_180 | Run3 | Barcode15 | T670G | 607 | 2 | no-detect |
| vssfru_233 | Run6 | Barcode53 | T670G | 580 | 9 | no-detect |
| vssfru_242 | Run6 | Barcode80 | T670G | 688 | 1 | no-detect |
| vssfru_245 | Run6 | Barcode89 | T670G | 769 | 6 | no-detect |
| vssfru_251 | Run7 | Barcode03 | T670G | 758 | 8 | no-detect |
| vssfru_261 | Run7 | Barcode15 | T670G | 764 | 6 | no-detect |
| vssfru_335 | Run9 | Barcode31 | C9866T | 385 | 2 | no-detect |

**Table S6.** Sequencing coverage of VUI-22JAN-01 samples with 1 mixed call (detect-mixed) at a variant defining position as defined by aln2type (Dataset 3).

| Sample ID | Run no. | Barcode | Position | Variant base | Sample-call | Status |
|---|---|---|---|---|---|---|
| vssfru_297 | Run8 | Barcode69 | A29510C | C | C,T | detect-mixed |
| vssfru_298 | Run8 | Barcode71 | A29510C | C | C,T | detect-mixed |
| vssfru_299 | Run8 | Barcode72 | A29510C | C | C,T | detect-mixed |
| vssfru_300 | Run8 | Barcode74 | A29510C | C | C,T | detect-mixed |
| vssfru_301 | Run8 | Barcode75 | A29510C | C | C,T | detect-mixed |
| vssfru_302 | Run8 | Barcode76 | A29510C | C | C,T | detect-mixed |
| vssfru_303 | Run8 | Barcode77 | A29510C | C | C,T | detect-mixed |
| vssfru_304 | Run8 | Barcode80 | A29510C | C | C,T | detect-mixed |
| vssfru_305 | Run8 | Barcode81 | A29510C | C | C,T | detect-mixed |
| vssfru_306 | Run8 | Barcode82 | A29510C | C | C,T | detect-mixed |
